# Whole Genome Sequencing in Premature Coronary Artery Disease in South Asians: A Pilot Case Control Study

**DOI:** 10.64898/2025.12.17.25342163

**Authors:** Iftikhar Ali Ch, Azhar Chaudhry, Fazal Jalil, Yasir Ali, Waseem Iqbal, Yusra Javed, Salman Khalid, Azeen Razzaq, Muhammad Azhar, Amna Nadeem, Tayyab Afzal, Naeem Tahirkheli, Ankur Kalra, Khurram Nasir

## Abstract

**Background/Objectives:** Coronary artery disease (CAD) remains the leading cause of mortality worldwide, with South Asia bearing a disproportionately high and rising burden, particularly at younger ages. This pilot study aimed to investigate genetic variants associated with premature coronary artery disease (PCAD) using whole genome sequencing (WGS).

**Methods:** WGS was conducted on 12 people (5 PCAD cases, 7 matched controls) to assess feasibility and methodology for future large-scale research. High-quality genomic DNA was sequenced at a minimum read depth of 10× with a quality threshold of Q30. Variant calling with stringent quality control identified single nucleotide polymorphisms (SNPs), followed by annotation against gnomAD for allele frequencies and ClinVar for pathogenicity. Protein-coding variants were filtered, and candidate genes were prioritized for comparative analysis between cases and controls.

**Results:** An average of over 8.8 million SNPs per individual was identified, with comparable overall variant distributions between cases and controls. Initial analyses revealed 120 SNPs exclusively present in PCAD cases. All protein-coding variants were rare (allele frequency <0.0001), and none were previously classified as pathogenic in ClinVar. After filtration, 87 candidate genes were prioritized. Enriched or unique variants in PCAD cases are mapped to genes involved in lipid metabolism, endothelial dysfunction, inflammatory signaling, immune regulation, thrombosis, vascular remodeling, and metabolic processes. Additional variants were identified in genes related to smooth muscle proliferation, oxidative stress, and other biological pathways.

**Conclusions:** This WGS pilot study provides an initial overview of the genomic landscape of PCAD in a South Asian cohort, highlighting potential rare variants across multiple biological pathways implicated in atherosclerosis that needs validation in a large-scale study.

**Graphical Abstract:** 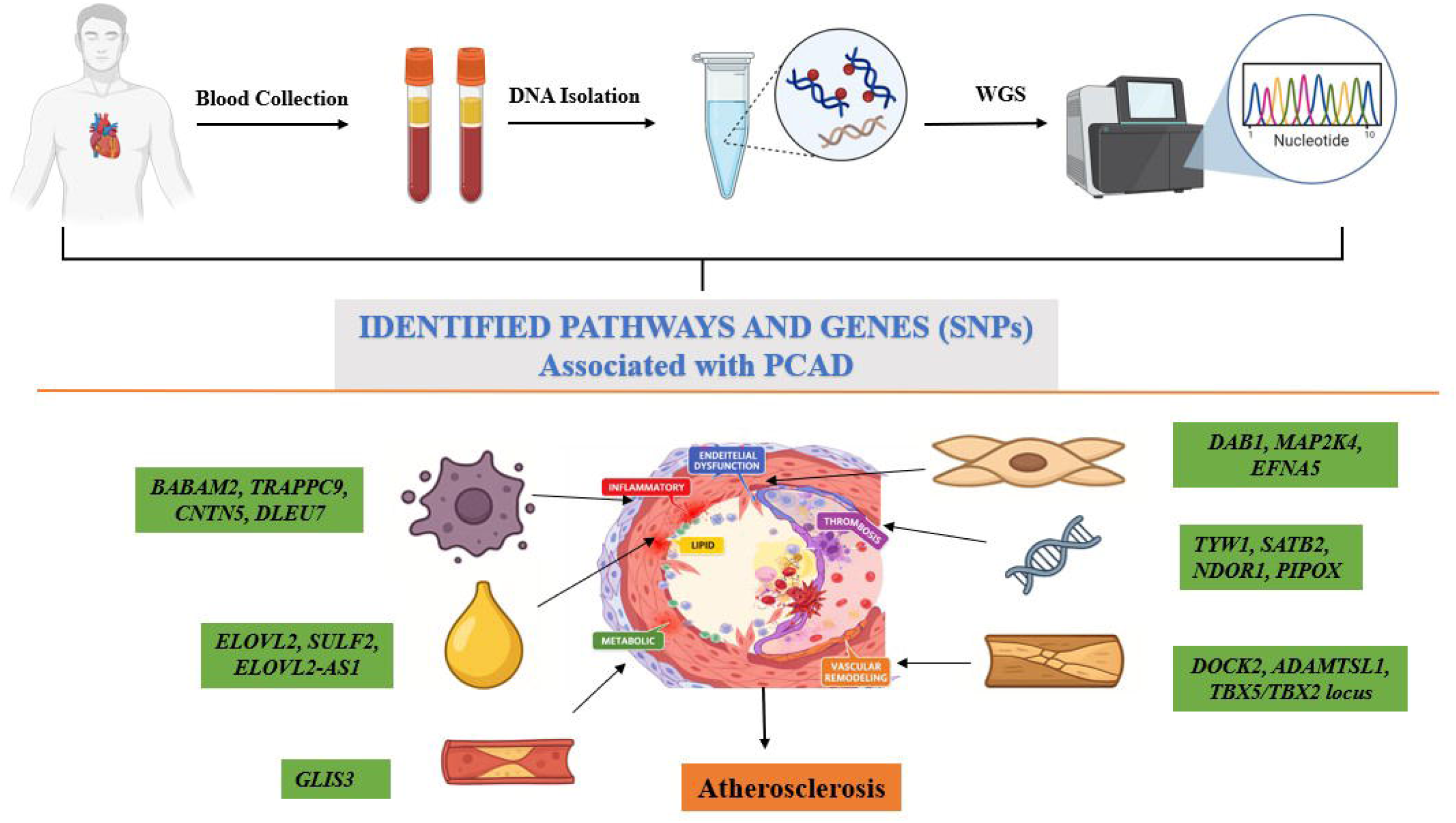

## 1. Introduction

Coronary artery disease (CAD) remains the leading cause of mortality worldwide, with South Asia experiencing a disproportionately high and escalating burden. According to the Global Burden of Disease (GBD) study, ischemic heart disease accounted for approximately 8.92 million deaths in 2015, underscoring its global health impact [1]. Of particular concern is the increasing incidence of premature CAD (PCAD), clinically defined as major cardiac events occurring before the age of 45 in men and 55 in women, with a more severe form affecting individuals under 40 years [2]. Epidemiological data of myocardial infarction (MI) cases reported that 21% of affected individuals were under 40, of which 79.8% were male, emphasizing a significant sex disparity and disproportionate impact on young men [3]. Notably, the incidence of acute MI demonstrates a marked age-dependent increase, from 2.1 per 100,000 person-years in individuals aged 20–29 to 16.9 in those aged 30–39 [4]. In Pakistan, the burden of cardiovascular disease (CVD) is escalating rapidly. Decades of industrial and economic impoverishment alongside significant challenges, including regional conflicts, natural disasters, and underinvestment in healthcare. Consequently, while infectious diseases remain prevalent, the healthcare system is now facing a growing crisis of non-communicable diseases, particularly CVD. According to the 2019 GBD study, Pakistan’s age-standardized incidence of CVD was 918.18 per 100,000 people (compared with the global rate of 684.33), and age-standardized death rate was 357.88 per 100,000 (compared to 239.85 globally) [5]. Additionally, data from the National Socioeconomic Registry Survey, comprising 34 million households, showed that 18.9% of participants self-reported having CVD [4] [5].

Globally, individuals of Indo-Asian origin exhibit the highest risk of developing CAD [6], hence establishing it as the leading cause of death in the Indo-Pakistani subcontinent. A national registry (2016–2019) analyzing percutaneous coronary intervention (PCI) data from 15,106 patients revealed that 52.6% had premature CAD (men aged 40–54; women aged 40–64), while 7.4% had extremely premature CAD (under 40 years) [7]. These statistics highlight a significant disease prevalence in younger populations, necessitating early interventions and preventive strategies.

The risk factors associated with PCAD differ across various ethnicities. Among Black populations, dyslipidemia, diabetes, and smoking are significant contributors, whereas in Whites, smoking is the primary factor. Among Hispanic populations, dyslipidemia, male sex, and family history are most prominent. Interestingly, no independent predictor has been found for Asian Indians [5]. Among South Asians, family history is a key contributor, warranting deeper investigation into genetic predisposition, including gene variants related to lipid metabolism, vascular function, and coagulation pathways [8] [9]. The elevated prevalence of consanguinity within the South Asian population likely increases the risk of homozygous mutations, contributing to early-onset and more aggressive disease. This is supported by epidemiological data reporting a positive family history in 18% of PCAD and 23% of early PCAD cases, suggesting a substantial genetic or familial environmental clustering [7]. In Pakistan, high age-adjusted prevalence of diabetes (30.8%), hypertension (37% in adults), and tobacco use (∼25% in adult males), coupled with a high consanguinity rate (58% first- or second-cousin marriages), synergistically amplify genetic risk [5].Multiple studies have explored genetic contributors to CVD in South Asians, including PROMIS (Pakistan), BRAVE (Bangladesh), LOLIPOP (UK), and the START cohort (Canada) [10] [11] [12] [13]. These studies, along with the 1000 Genomes Project [14], have provided critical insight into gene-environment and gene-gene interactions. The C4D Genetics Consortium meta-analysis of four GWAS (PROMIS, LOLIPOP, PROCARDIS, and HPS) identified 59 single nucleotide polymorphisms (SNPs) linked to CAD, five achieved genome-wide significance, implicating genes involved in lipid metabolism, inflammation, and vascular remodeling [15]. Furthermore, a multiethnic GWAS meta-analysis combining seven non-South Asian and two South Asian cohorts identified 25 CAD-associated SNPs within or proximal to genes such as PECAM1 (inflammation), LMOD1 (vascular smooth muscle), and PROCR (coagulation) [16]. PROMIS study also identified six novel SNPs in the CX3CR1 gene specific to South Asians that were not reported in larger European studies, suggesting ethnicity-specific genetic architecture [17]. Another study in young South African Indians reported an association between the p53 Arg72 allele and PCAD, proposing it as a potential genetic marker for early disease [18]. Epigenetic research further supports the gene-environment interplay whereby environmental exposures (e.g., diet, maternal health) modulate disease risk through gene regulation, as evident by the emergence of miR-21 as a potential epigenetic marker for atherogenic dyslipidemia in South Asians [19].

Although significant discoveries have been made in middle-aged and older individuals, the genetic factors influencing PCAD in younger populations—particularly within understudied regions like Indo-Pakistan—are still not well understood. This highlights the need for high-throughput whole genome sequencing (WGS) to identify variants specific to PCAD. Our research is designed to address an important gap by launching a pilot study, which could lead to larger studies focused on discovering new mutations, SNPs, and structural variants in PCAD. This initial study may help lay the foundation for creating polygenic scores in broader research, ultimately enhancing early diagnosis, risk assessment, and tailored treatments (20,21).

## 2. Methodology

### 2.1 Study Population

This retrospective case-control pilot study explores WGS as a method for investigating PCAD. The study enrolled 12 participants, including 5 PCAD patients (PCAD group) and 7 matching controls (control group). Blood samples were collected from Armed Forces Institute of Cardiology/ National Institute of Heart diseases, Rawalpindi, Pakistan. The inclusion criteria for patient selection were as follows: (1) clinical symptoms indicative of myocardial infarction (MI) lasting over 20 minutes within the previous 24 hours of preceding index hospitalization, (2) ECG changes consistent with MI, such as new pathologic Q waves, at least 1mm ST elevation in two or more contiguous limb leads, a new left bundle branch block, or persistent ST-T wave changes diagnostic of non-Q wave MI, and (3) those with elevated troponin-T levels.

Exclusion criteria included recent or active infection within the past 2 weeks, chronic infections (e.g., HIV, hepatitis B, hepatitis C, tuberculosis), chronic autoimmune diseases (e.g., rheumatoid arthritis, lupus), acute pericarditis, end-stage renal failure, history of malignancy, and those who refuse to provide consent. Individuals in the control groups were matched to cases by age, gender and co-morbidities (within 5-year age bands) without a history of cardiovascular disease.

### 2.2 Sample Collection

A blood sample (5mL) was collected from each study participant under the supervision of a certified physician. During sample collection, baseline characteristics and clinical information were documented using a specifically designed questionnaire. After collection, blood samples were immediately transported under controlled conditions to the Chugtai laboratory Jail Road Lahore, Pakistan and stored at 4°C until further processing.

### 2.3 DNA Extraction and Analysis

Genomic DNA was isolated from whole blood using the standard organic phenol-chloroform method, following the manufacturer’s instructions (Sambrook, 1989). The extracted DNA was quantified and stored at 4°C for subsequent experimentation.

### 2.4 Sample Quality Control

DNA integrity was assessed by electrophoresis on a 1% agarose gel. Initial quantification analysis was performed using a NanoPhotometer spectrophotometer (Implen GmbH), and DNA concentration and purity were subsequently confirmed with a Qubit 2.0 Fluorometer (Thermo Fisher Scientific).

### 2.5 Library Preparation

Genomic DNA was sheared to ∼350 bp fragments using an ultrasonic processor. The resulting fragments were then used to construct sequencing libraries through a standardized workflow, including end repair, adenylation at the 3′ ends, adapter ligation, purification, and polymerase chain reaction (PCR) amplification. The resulting sequencing libraries were quantified using a Qubit 2.0 Fluorometer and samples were normalized to a working concentration of 1 ng/µL. Library fragment size distribution was verified using an Agilent 2100 Bioanalyzer, and final library concentrations were confirmed by quantitative PCR prior to sequencing.

### 2.6 Whole Genome Sequencing

WGS was performed on an Illumina NovaSeq 6000 (paired-end 150 bp) platform using S4 flow cell. FASTQ formatted files were received from the sequencing provider. Sequencing quality was checked using FastQC, which includes (1) more than 50Gbp bases in each FASTQ file (each sample have two paired FASTQ files), (2) 150bp length for all sequences, and (3) pass all the quality check terms in FastQC report, which include per base sequence quality, per sequence GC content, sequence length distribution, sequence duplication levels, overrepresented sequences, adapter content. Genome Reference Consortium Human Build 38 (GRCh38) reference panel provided by Genomic Analysis ToolKit (GATK) was chosen as a reference genome. Read alignment and variant calling were performed using the graphics processing unit (GPU) accelerated GATK pipeline in Nvidia Clara Parabricks v4.3.1. After receiving GVCF files for each sample, joint genotyping was performed using GLnexus v1.2.6 to convert GVCF files into a multi-sample VCF file.

### 2.7 Bayesian Model for Detecting Associations (BMDA) SNP Analysis Pipeline

This analysis pipeline started with the VCF file generated after WGS. The GFF (General Feature Format) or GTF (Gene Transfer Format) file was used for reference genome. BMDA analysis included the following steps:

1) Variant Filtering:

The raw variant call format (VCF) file was filtered using “vcftools” using a minimum quality score of 30, and minimum depth of coverage of 10 reads.

2) Basic VCF statistics:

Program “vcf-stats” was used to extract the numbers of each SNP for each sample. Average ts/tv (transition SNPs to transversion SNPs) was 1.99.

3) Extracting the initial SNP report:

An in-house script was used to extract the initial report. The report was generated where only rows containing control samples with identical genotypes and patients with different genotypes were picked.

4) Preparing GFF and GTF file:

An annotation table was generated from the GTF annotation file to extract the relevant information.

5) SNP Annotation:

The initial SNP report from step 3 was amended with the gene annotation.

### 2.8 Variants annotation and filtering

Sequence alignment to the GRCh38 (hg38) reference genome was performed using the Burrows-Wheeler Aligner (BWA version 0.7.15), followed by duplicate removal with Picard. Subsequent processing, including indel realignment, base quality score recalibration, and variant calling, was conducted with the GATK. ANNOVAR (v2019Oct24) was used for variant annotation. Allelic frequencies were filtered and verified using public genomic databases, including the Genome Aggregation Database (gnomAD), Kaviar Genomic Variant Database, the Greater Middle East (GME) Variome Project, and the 1000 Genomes Project (1000G).

**Figure 1.**
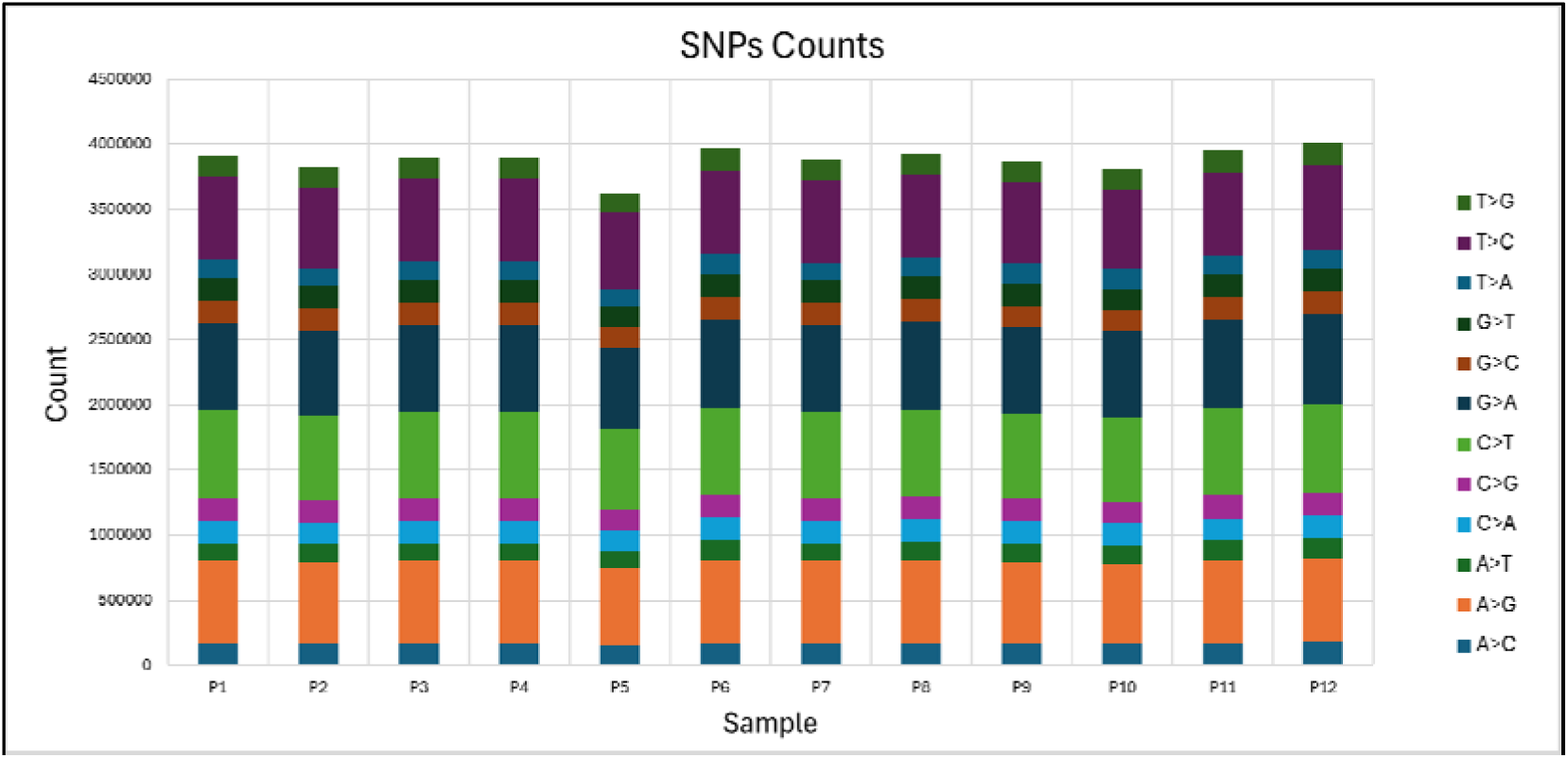
Distribution of Single Nucleotide Polymorphism (SNP) Substitution Types Across 12 Samples. The stacked bar chart shows the counts of different SNP substitution types (e.g., A>G, C>T, T>C, etc.) across twelve whole genome sequencing samples (P1–P12). Each colored segment represents a specific nucleotide substitution, with transitions (e.g., A>G, C>T) and transversions (e.g., A>T, C>G) clearly visualized. The total SNP burden is comparable across samples, indicating consistent sequencing depth and variant calling quality. The most common substitutions include C>T and G>A, which are known transition mutations typically resulting from deamination events.

**Figure 2.**
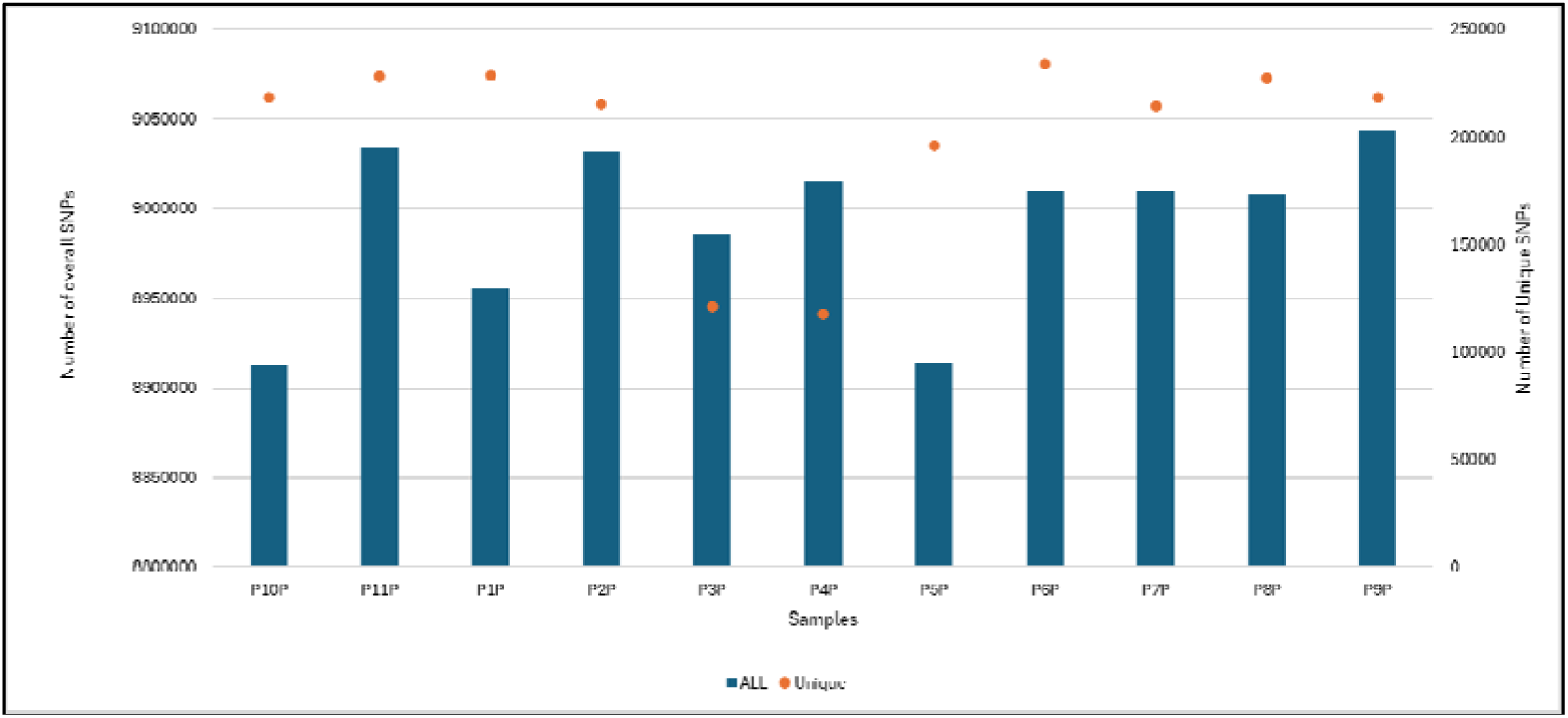
Comparison of Overall and Unique SNP Counts Across Whole Genome Sequencing Samples. The bar and dot plot illustrate the total number of single nucleotide polymorphisms (SNPs) (blue bars, left y-axis) and unique SNPs (orange dots, right y-axis) for each individual sample (P1P–P11P). While the total SNP counts are relatively consistent across all samples, unique SNP counts show some inter-sample variability, suggesting potential individual-specific or population-specific variations. This comparison highlights both the sequencing depth and the uniqueness of genetic variation within each genome in the context of premature coronary artery disease analysis.

## 3.0 Results

### 3.1 Patient characteristics

This pilot study comprised a cohort of 12 participants (5 PCAD cases, 7 controls). Cases were modestly older than controls (mean age 39.0 ± 3.4 vs 34.4 ± 6.3 years). The cohort was predominantly male (cases= 4 of 5, controls= 6 of 7). All participants were recruited from the northern regions of Pakistan, including Rawalpindi, Swat, and Mardan. Adiposity indices trended higher among cases (mean BMI: 28.9 ± 6.4 kg/m² vs 25.1 ± 4.7 kg/m², weight: 81.7 ± 8.0 kg vs 72.9 ± 14.5 kg). Hypertension and dyslipidemia were documented in one case whereas no participant, in either group, reported diabetes mellitus, smoking history, chronic kidney disease, prior MI/PCI/CABG, or family history of CAD. Only 2 of 5 cases were on statin therapy prior to MI. Baseline medications listed in Table1.

### 3.2 Laboratory values

Lipid profiles were broadly similar between groups, with substantial variability. Mean total cholesterol was 163.0 ± 73.8 mg/dL in cases vs 174.0 ± 30.9 mg/dL in controls; LDL-C 87.8 ± 71.6 vs 125.8 ± 22.8 mg/dL; HDL-C 38.5 ± 4.5 vs 35.6 ± 3.0 mg/dL; and triglycerides 186.2 ± 87.2 vs 178.2 ± 61.2 mg/dL. HbA1c averaged 5.8 ± 0.5% in cases and 5.4 ± 0.1% in controls.

Serum creatinine was modestly higher among cases (1.1 ± 0.2 vs 0.8 ± 0.1 mg/dL). Data were presented as mean ± SD or count ‘n’ (%).

### 3.3 Genetic testing results

WGS was performed on a cohort of 12 individuals to investigate potential genetic variants associated with PCAD. The cohort comprised 7 control individuals (samples P1P to P7P) and 5 confirmed PCAD cases (samples P8P to P12P). High-quality DNA libraries were sequenced with a minimum read depth of 10x and a quality score threshold of 30. Following variant calling and quality control, over 8.8 million SNPs were detected per sample, with total SNP counts comparable across all individuals. Unique SNPs, representing potentially disease-associated or population-specific variations, were quantified separately and exhibited slight variability within the PCAD group. We identified 120 SNPs exclusively present in patients. All protein-coding variants annotated against gnomAD were rare (allele frequency < 0.0001) and none were previously classified as pathogenic in ClinVar. These variants were annotated and filtered for protein-coding regions, yielding 87 candidate genes (full list is presented in Supplementary Data S1).

Comparative analyses identified SNPs uniquely present or significantly enriched in PCAD cases, mapped to genes involved in atherosclerosis, lipid metabolism, inflammation, and endothelial dysfunction. Several notable SNPs were identified in genes implicated in diverse pathophysiological pathways associated with atherosclerosis, including lipid metabolism (*ELOVL2, SUGP1, SULF2*), endothelial dysfunction (*ARAP2, EFNA5 MAP2K4, NABP,, POR, RAP1GAP*), inflammatory signaling (*BABAM2, CD28, CNTN5, DLEU7, HDAC9, ITPR1, POR, PLCB1, TSPAN33, TRAPPC9*), immune regulation (*DLEU7, IL4I1, KIR2DS4*), thrombosis (*MAP2K4, PLCB1, RAP1GAP, SULF2,TSPAN33,*), vascular remodeling (*ADAMTSL1, BCAS3, DOCK2, ITGA8, TBX5,*), and metabolic processes (*ARHGAP44,GLIS3*). Additional variants were observed in genes linked to smooth muscle proliferation, oxidative stress or other processes *(ACAN, CDK14, LRP2, NDOR1, PIPOX, RUBCN, SATB2, TUBB4B, TYW1)*.

The *LINC01744* gene exhibited the largest number of variants (n=19), followed by *ELOVL2* with (n=14). *DAB1* and *TRERNA1* each showed 10 variants, while *DLEU7*, *KNTC1*, and *TPT1-AS1* had 9 each. *MAP2K4* carried 8 variants, and *ELOVL2-AS1* and *KCNIP4* had 7 variants each.

**Figure 3.**
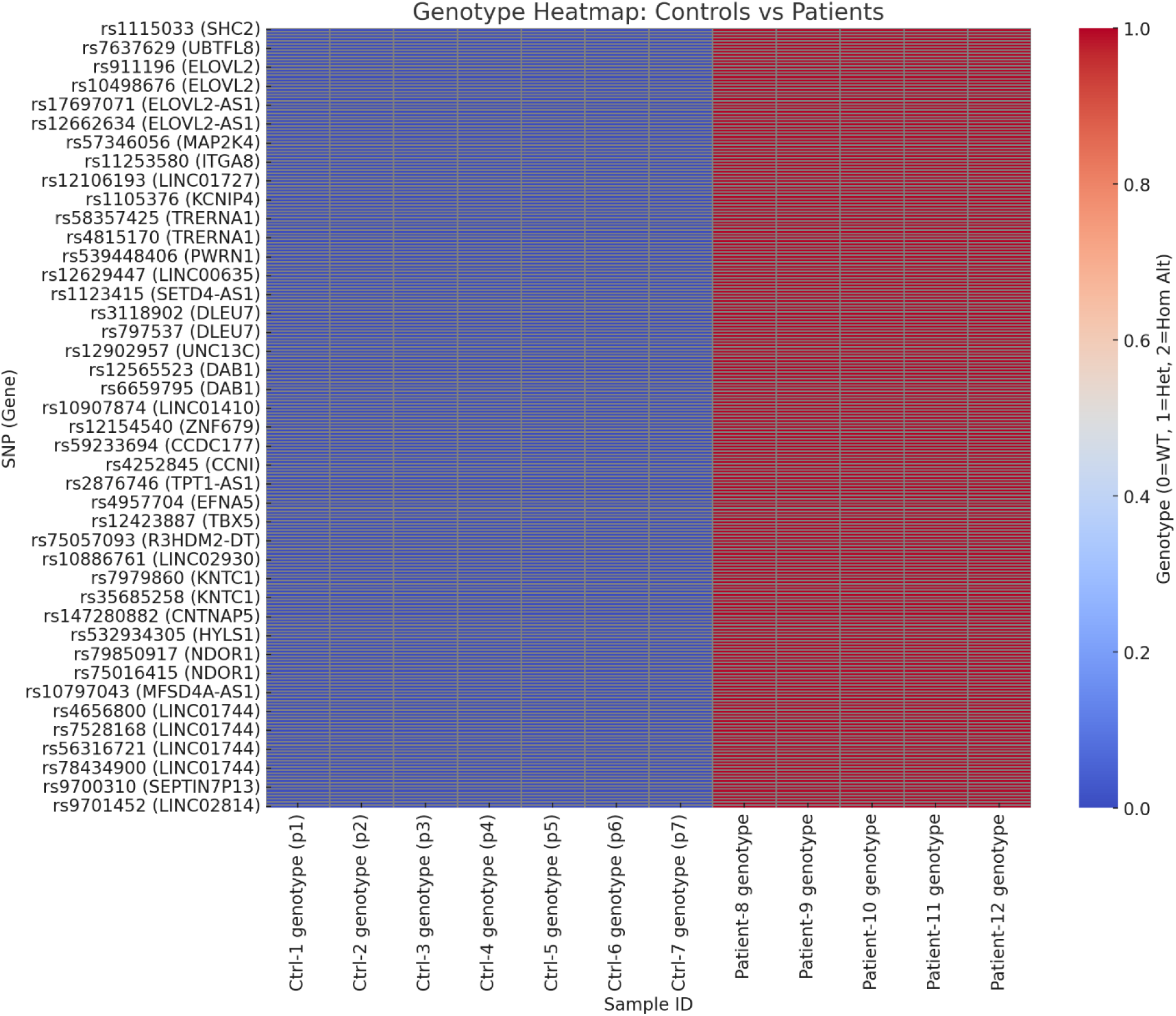
This heatmap visualizes the genotypic variation across 40 selected SNPs (y-axis), each labeled with its corresponding gene, in 12 study participants (x-axis). Samples Ctrl-1 to Ctrl-7 represent healthy controls, while patient-8 to patient-12 represent individuals with premature coronary artery disease (PCAD). A genotype heatma shows the differences between controls and patients across all disease-associated SNPs. **Blue (0)**: Homozygous reference (0|0) – observed in all controls. **Light to dark red (1 or 2)**: Variant genotypes – seen only in patients. **Consistent red bands** across patient columns highlight recurrent SNPs potentially contributing to **PCAD**.

**Figure 4.**
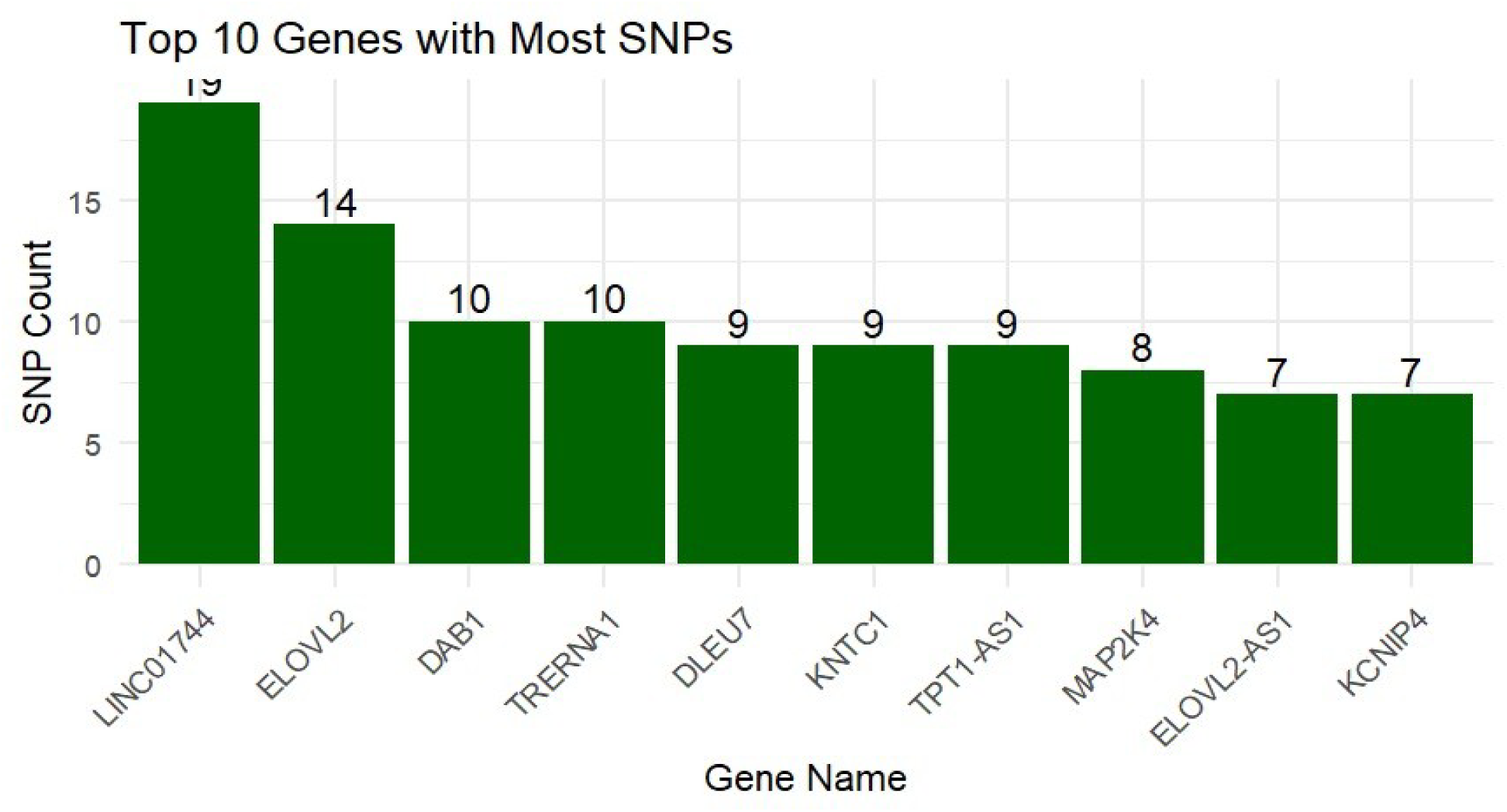
Bar chart illustrating the ten genes with the greatest number of SNPs identified across the WGS dataset with LINC01744 exhibiting the highest SNP burden (19 variants), followed by ELOVL2 (14 SNPs), DAB1 and TRERNA1 (10 each), DLEU7, KNTC1, and TPT1-AS1 (9 each). Other notable genes include MAP2K4 (8), ELOVL2-AS1 (7), and KCNIP4 (7).

**Figure 5.**
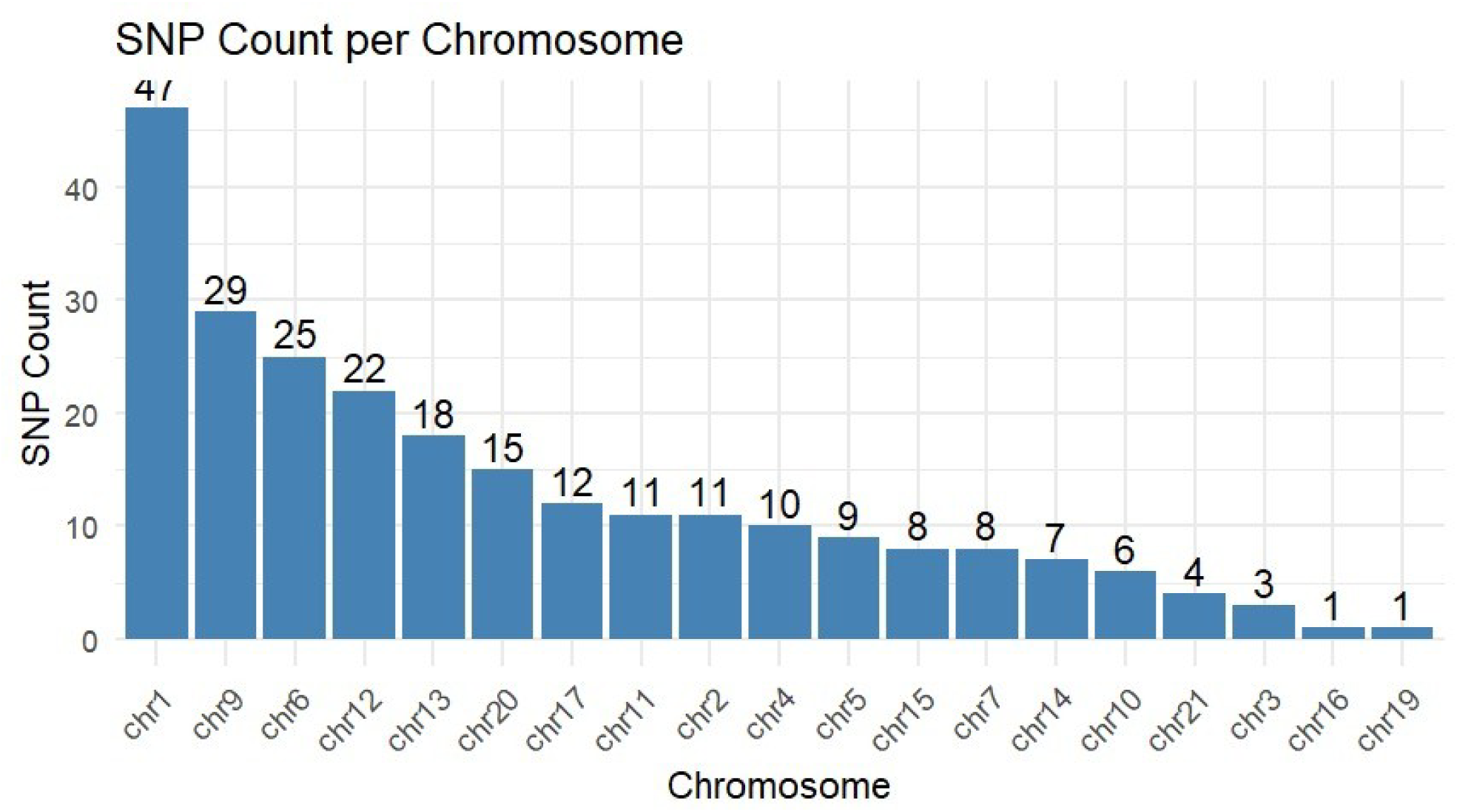
Bar graph showing SNPs count per chromosome from WGS data, with Chromosome 1 exhibiting the highest SNP burden of variants (47), followed by chromosome 9 (29), chromosome 6 (25), and chromosome 12 (22). Chromosomes 3, 16, and 19 had the fewest SNPs (1 each).

## 4.0 Discussion

### Functional relevance of genes with SNPs

From a group of 12 subjects (5 with PCAD, 7 healthy), we found 120 SNPs unique to PCAD cases. Protein-coding variants (n = 87 unique genes) underwent allele frequency annotation using gnomAD, all were rare (AF < 0.0001), and none were previously classified as pathogenic in ClinVar. A total of 33 SNPs were identified in non-coding regions and are detailed in the supplementary material.

This summary outlines the reported SNPs and their potential biological pathways linked to atherosclerosis. The supplementary material provides more in-depth associations and mechanisms with PCAD. It’s important to note that, as an exploratory pilot study, our research does not establish causality. Instead, these findings serve as groundwork for future, larger-scale studies aimed at validation.

### 4.1 Lipid metabolism

Dysregulation of lipid metabolism represents a central mechanistic axis in the pathogenesis of PCAD. Beyond traditional lipid measures, genetic and molecular perturbations in **cholesterol transport, lipoprotein remodeling, lipid oxidation, and intracellular lipid handling** drive early atherosclerosis. Genetic Variants observed in this study like ELOVL2 and ELOVL2-AS1 may affect PUFA remodeling, LDL processing, and lipoprotein clearance, influencing vascular inflammation and plaque development (22,23,24). Case-restricted heterozygosity across multiple rsIDs suggests a locus-level burden rather than a single causal SNP—very consistent with PUFA-pathway biology. ELOVL2 plays a key role in PUFA metabolism and lipid homeostasis, making it relevant for PCAD patients. Likewise, SUGP1 and SULF2 variants are linked to cholesterol metabolism, reduced triglyceride clearance, and CAD (25,26) (Table 1). Investigating genetic variants governing these pathways in large scale studies would help validate these findings and guide risk prediction, early detection and potentially novel treatment strategies. Developing functional assays in cellular models could aid in prioritizing candidates for therapeutic intervention; however, these findings must first be validated through adequately powered studies.

**Table 1.**
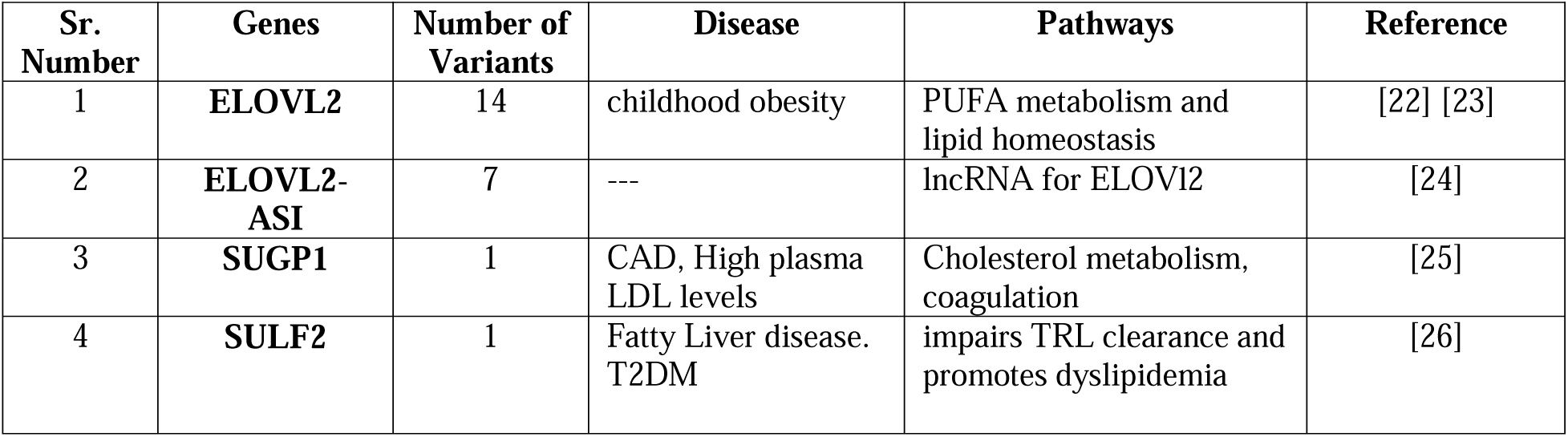
Variants in lipid-metabolism-related genes.

| Sr. Number | Genes | Number of Variants | Disease | Pathways | Reference |
| --- | --- | --- | --- | --- | --- |
| 1 | ELOVL2 | 14 | childhood obesity | PUFA metabolism and lipid homeostasis | [22] [23] |
| 2 | ELOVL2-ASI | 7 | --- | lncRNA for ELOVL2 | [24] |
| 3 | SUGP1 | 1 | CAD, High plasma LDL levels | Cholesterol metabolism, coagulation | [25] |
| 4 | SULF2 | 1 | Fatty Liver disease. T2DM | impairs TRL clearance and promotes dyslipidemia | [26] |

**Figure 6.**
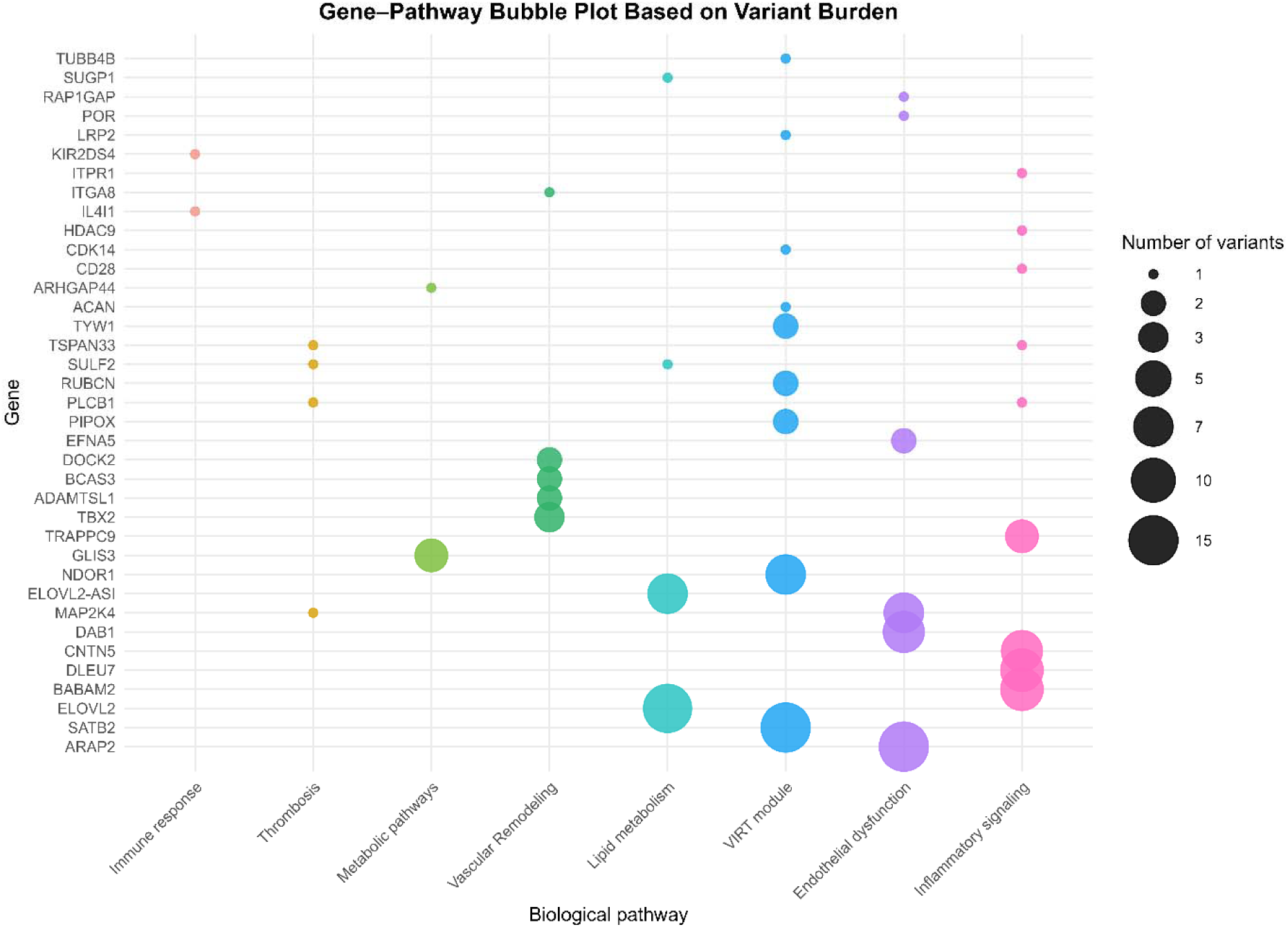
Bubble plot showing the distribution of genetic variants on biological pathways of cardiac-related genes. Each bubble represents a gene annotated to a particular biological pathway. The size of the bubble shows the variants per gene, and the colors denote the pathways, such as lipid metabolism, endothelial dysfunction, inflammatory signaling, immune response, thrombosis, vascular remodeling, metabolic pathways, and VIRT module.

### Endothelial cell function

Endothelial cells orchestrate vascular homeostasis by regulating nitric oxide (NO)-mediated vasodilation, antithrombotic surface properties, barrier integrity, and leukocyte trafficking. Several genetic variants identified in the pilot study are hypothesized to have mechanistic relevance in the development of premature atherosclerosis. Specifically, DAB1 may contribute through activation of the Reelin pathway (27,28,29); ARAP2 and MAP2K4 may be involved via leukocyte adhesion (30,31,32); and EFNA5, POR, and RAP1GAP may influence disease progression through mechanisms related to vascular remodeling and nitric oxide release (33–39). Exposure to pathological stimuli, such as disturbed blood flow, hyperglycemia, dyslipidemia, tobacco exposure, or oxidative stress, induces endothelial dysfunction leading to reduced eNOS activity and NO bioavailability, elevated reactive oxygen species (ROS) driving redox-sensitive pathways (NF-κB/JNK); induction of cell surface adhesion molecules (ICAM-1, VCAM-1, E-selectin), resulting in diminished barrier function leading to increased permeability to inflammatory mediators (monocytes, and apoB lipoproteins). In this pathophysiological framework, it is proposed that these variants converge on key processes including endothelial signaling, redox homeostasis, cellular adhesion, and tissue repair, thus representing biologically plausible mechanisms linking genotype to early atherosclerosis, irrespective of prolonged pathological stimuli. While the current pilot study lacks sufficient statistical power to demonstrate causality, it highlights the need for extensive studies to confirm these preliminary observations (Table 2).

**Table 2.** Variants identified in EC-function-related genes.

| Sr. Number | Genes | Number of Variants | Disease | Pathways | Reference |
| --- | --- | --- | --- | --- | --- |
| 1 | **DAB1** (Disabled-1) | 8 | | Activation of the Reelin pathway activates NF-κB, increases expression of ICAM-1, VCAM, and E-selectin | [27], [28] [29] |
| 2 | **ARAP2** (Arf/Rho GAP) | 15 |  | Integrin signaling, leukocyte–endothelial adhesion | [30], [31] |
| 3 | **MAP2K4** | 7 |  | Activator of JNK, promotes endothelial apoptosis and pro-inflammatory gene expression | [32] |
| 4 | **EFNA5** (Ephrin-A5) | 2 | Cardiovascular development, Atherosclerosis | Monocyte adhesion via EPHA2/EPHA4 and RhoA-cytoskeletal remodeling, Ca<sup>2+</sup>/NFAT pathway | [33] [34] |
| 5 | **POR** (NADPH–cytochrome P450 oxidoreductase) | 1 |  | eNOS/AKT signaling, NO bioavailability, EC dysfunction. | [35] [36] [37] |
| 6 | **RAP1GAP** Rap1 GTPase-activating protein | 1 | myocardial infarction (MI) | Endothelial NO release, Worsen MI via the AMPK/SIRT1/NF-κB pathway | [38] [39] |

### 4.2 Inflammatory pathways

Atherosclerosis is a chronic inflammatory disease characterized by endothelial dysfunction which leads to subendothelial retention of lipoproteins, activation of adhesion molecules (VCAM-1, ICAM-1, E-selectin), and chemokines resulting in the recruitment of monocytes and T cells into the intima. SNPs found in the pilot study, such as BABAM2, DLEU7, CNTN5, and TRAPPC9, may affect NF-κB signaling or adhesion proteins, promoting inflammation (40-44). These variants can disrupt innate immune pathways (TLR–MyD88–NF-κB, NLRP3–IL-1β), leading to macrophage activation, foam cell formation, and reduced efferocytosis. Additional genes with single variants, HDAC9, CD28, ITPR1, TSPAN33 and PLCB1, might also contribute to a pro-inflammatory state by either increasing inflammation or reducing protective mechanisms (50-59). Details of these variants are provided in Table 3. These findings suggest that investigating genetic variants and their underlying mechanisms—especially those responsible for inflammation—could hypothetically be an effective approach to further our understanding of premature conditions.

**Table 3.** Variants identified in inflammatory pathway-related genes.

| Sr. Number | Genes | Number of Variants | Disease | Pathways | Reference |
| --- | --- | --- | --- | --- | --- |
| 1 | **BABAM2**<br>BRISC/BRCA1-A complex mem2 | 9 | Acute myocardial infarction, | Intersects TNF/NF-κB and cell-death/DNA-damage axes | [40] [41] [42] |
| 2 | **DLEU7** | 9 | Cardiovascular Diseases | inhibits the B-cell receptors, dampening NF-κB/NFAT signaling | [43] [44] |
| 3 | **CNTN5** | 8 |  | neuronal adhesion protein | [45] [46] [47] [48] |
| 4 | **TRAPPC9** | 4 |  | Promotes NF-κB signaling | [49] |
| 5 | **HDAC9** | 1 | CAD Large artery ischemic stroke | Increasing pro-inflammatory signaling via IKK/NF-κB | [50] [51] |
| 6 | **CD28** | 1 | Acute coronary syndrome (ACS), Rheumatoid arthritis (RA) | Expression on Tregs supports immune regulation and appears protective | [52] [53] |
| 7 | **ITPR1**<br>(IP<sub>3</sub> receptor 1) | 1 | CAD | IP<sub>3</sub>R-mediated Ca<sup>2+</sup> release, NO signaling, VSMCs contractility, phenotype switching | [54] [55] [56] |
| 8 | **TSPAN33** | 1 |  | macrophage activation via NOTCH / TLR | [57] |
| 9 | **PLCB1** | 1 | Coronary artery aneurysm (CAA) Kawasaki disease | Reduce endothelial cell inflammation, Platelet activation pathways downstream of GPCR. | [58] [59] |

### 4.3 Immune-related pathways

Atherosclerosis is also postulated to have maladaptive immune response to retained apoB-lipoproteins within the arterial wall. Innate sensors (TLRs, scavenger receptors) on endothelial cells, macrophages, and dendritic cells detect modified LDL and cellular debris, activating NF-κB and inflammasomes (e.g., NLRP3→IL-1β/IL-18). This inflammatory milieu is perpetuated through monocyte recruitment, macrophage polarization, defective efferocytosis, and neutrophil extracellular traps, creating a non-resolving inflammatory cascade. In this pilot study we noted two variants, KIR2DS4 and IL4I1 that can hypothetically alter immune response by triggering immune response or suppressing protective mechanisms (60,61). Adaptive immunity layers amplify this niche wherein Th1- and Th17-derived cytokines (IFN-γ, IL-17) exacerbate vascular inflammation while Tregs counteract this progression via IL-10/TGF-β. B-cells exhibit a dichotomous response, with natural IgM against oxLDL being protective, whereas some B2 responses promote disease (Table 4). Natural killer (NK) cells and cytotoxic T cells can add plaque-destabilizing cytotoxicity. Genes that bias Treg/Th17 balance, NK activation thresholds, or cytokine set-points can therefore shift risk—offering handles for diagnostics (circulating proteins/cell-subset signatures) and therapeutics (pathway-targeted immunomodulation). Though purely hypothetical but it is an important avenue to explore in large sample studies with subsequent validation studies in cell and animal models.

**Table 4.** Variants identified in immune-related genes.

| Sr. Number | Genes | Number of Variants | Disease | Pathways | Reference |
| --- | --- | --- | --- | --- | --- |
| 1 | **KIR2DS4** | 1 | HIV | **Reduce CD4+ T-cell counts** in chronic HIV-1 | [60] |
| 2 | **IL4I1**<br>(interleukin-4–induced-1) | 1 |  | Promote regulatory T cells (Tregs). Suppress pro-inflammatory Th17 cells | [61] |

### 4.4 Thrombotic pathways

MI represents the clinical manifestation of ruptured atherosclerotic plaque exposing tissue factor and subendothelial matrix, leading to a cascade of platelet activation and coagulation, which culminates in occlusive coronary thrombosis. Genetic predispositions and acquired risk factors that heighten platelet reactivity, enhance thrombin generation, or impair endothelial anticoagulation mechanisms alter this balance toward occlusive arterial thrombosis. Several genetic variants identified in this preliminary study may suggest an increased thrombotic risk, specifically involving PLCB1, MAP2K4, and SULF2 (58,62). These variants could contribute to early-onset myocardial infarction by activating thrombogenic pathways, even in cases where coronary atherosclerosis is minimal. The variants associated with thrombotic pathways are detailed in Table 5.

**Table 5.** Variants associated with thrombotic pathways.

| Sr. Number | Genes | Number of Variants | Disease | Pathways | Reference |
| --- | --- | --- | --- | --- | --- |
| 1 | **PLCB1** | 1 |  | Platelet Ca<sup>2+</sup> signaling granule release | [58] |
| 2 | **TSPAN33** | 1 | --- | increases ADAM10 activity | ---- |
| 3 | **MAP2K4** | 1 |  | Lower the threshold for secretion, stabilize thrombi |  |
| 4 | **SULF2** | 1 |  | Reducing local AT/TFPI efficacy | [62] |

### 4.5 Vascular remodeling

Vascular remodeling is the structural and cellular reconfiguration of the arterial wall in response to hemodynamic load, injury, and inflammation. It encompasses endothelial activation, smooth-muscle cell (SMC) phenotypic switching (contractile to synthetic/migratory), extracellular-matrix reorganization, microfibril/TGF-β signaling, integrin-mediated transduction, and (neo)angiogenesis within the intima and media. The study identified variants in the DOCK2, ADAMTSL1, and TBX2 genes (63–70) as potential modulators of signaling pathways. Additional variants, such as ITGA8, are considered to influence the composition of the extracellular matrix (71,72), while BCAS3 is implicated in the regulation of angiogenesis (73,74). Table 6 provides an overview of these variants and their associated diseases.

**Table 6.** Variants identified in vascular remodeling-associated genes.

| Sr. Number | Genes | Number of Variants | Disease | Pathways | Reference |
| --- | --- | --- | --- | --- | --- |
| 1 | **DOCK2** | 2 | Myocarditis, cardiac graft rejection | Knockdown blunts TNF-α-induced ICAM-1/VCAM-1/MCP-1 and NF-κB activation. | [63] [64] [65] |
| 2 | **ADAMTSL1** | 2 | | Binds fibrillin-1, modulates TGF-β bioavailability. | [66] |
| 3 | **TBX2** | 3 | Cardiovascular malformations | Developmental transcription factor, linked to cardiac electrical traits and AF. | [67] [68] [69] [70] |
| 4 | **ITGA8** | 1 |  | Regulates differentiation, migration, and ECM/fibrotic responses. | [71], [72] |
| 5 | **BCAS3** | 2 | Coronary artery disease | Angiogenesis and vascular remodeling activate CDC42 and the actin cytoskeleton. | [73] [74] |

While adaptive remodeling preserves lumen caliber, maladaptive remodeling (excess SMC migration/proliferation, aberrant extracellular matrix deposition, fibrous cap thinning, and plaque neovascularization) promotes plaque growth, instability, and thrombosis. Genetic variants that alter leukocyte–endothelial crosstalk, SMC adhesion/motility, matrix architecture, or endothelial polarity/angiogenesis can predispose an individual to maladaptive remodeling, thereby increasing susceptibility toward premature atherogenesis and MI. These mechanisms are outlined to generate hypotheses for future large-scale studies. The increasing incidence and earlier onset of PCAD call for focused research into these mechanisms.

### 4.6 Metabolic pathways

Metabolic syndrome, diabetes, and insulin resistance are potent atherogenic precursors because they create a lipoprotein-rich, pro-inflammatory, NO-deficient, and prothrombotic vascular environment, thereby transforming metabolic stress into endothelial dysfunction, plaque growth, and ultimately thrombotic coronary events. Genetic determinants of insulin resistance are of significant interest for advancing our understanding of mechanistic pathways leading to premature atherosclerosis. This study observed GLIS3 and ARHGAP44 variants that may be hypothetically connected to this pathway (75-77) (Table 7).

**Table 7.** Variants identified in metabolic pathway-related genes.

| Sr. Number | Genes | Number of Variants | Disease | Pathways | Reference |
| --- | --- | --- | --- | --- | --- |
| 1 | **GLIS3**<br>β-cell transcript factor | 4 | Insulinopenia, hyperglycemia, | adult β-cell function, insulin gene transcription | [75] [76] |
| 2 | **ARHGAP44** | 1 | Glucose metabolism and neuronal development | Associated with HbA$_{1c}$ levels | [77] |

### 4.7 Vascular Inflammation–Remodeling–Thrombosis (VIRT) Pathway in PCAD

The identified genes converge on key hypothetical mechanisms underlying atherosclerosis at young age, including vascular senescence, oxidative stress, inflammation, and plaque instability. Variants in SATB2 mapped to the 9p21/CDKN2A/B locus, implicating endothelial senescence and VSMC osteogenic programming via RUNX2 [78,79]. NDOR1 variants were linked to diabetic vascular aging through dysregulation of redox-sensitive NF-κB and MAPK pathways [80–82]. Metabolic and translational stress pathways were represented by TYW1 and PIPOX, involving tRNA modification and H O -generating amino-acid metabolism, respectively [83,85,86]. RUBCN variants, associated with acute coronary syndrome, impair LC3-associated phagocytosis and efferocytosis, contributing to plaque inflammation and vulnerability [84]. Additional genes (LRP2, CDK14, TUBB4B, ACAN) influence renin–angiotensin homeostasis, Wnt signaling, cytoskeletal-AKT pathways, and VSMC apoptosis, collectively reinforcing pathways of vascular dysfunction and plaque instability [87–90]. This pilot study revealed nine VIRT pathway-related genes harboring 32 variants that need further exploration in large study. These genes are summarized in Table 8.

**Table 8.** Variants identified in VIRT pathway-related genes.

| Sr. Number | Genes | Number of Variants | Disease | Pathways | Reference |
| --- | --- | --- | --- | --- | --- |
| 1 | **SATB2** | 15 |  | Endothelial senescence at 9p21/CDKN2A/B locus, VSMC osteogenic programming via RUNX2 | [78] [79] |
| 2 | **NDOR1**<br>NADPH-dependent diflavin oxidoreductase 1 | 7 | diabetic vascular aging | Affects redox-sensitive signaling pathways such as NF-κB and MAPKs | [80] [81] [82] |
| 3 | **TYW1** | 2 |  | Modifies tRNA<sup>Phe</sup> (wybutosine pathway) | [83] |
| 4 | **RUBCN**<br>(Rubicon) | 2 | acute coronary syndrome (ACS) | LC3-associated phagocytosis (LAP); impairs efferocytosis, | [84] |
| 5 | **PIPOX** | 2 |  | Glycine/sarcosine metabolism, generate H<sub>2</sub>O<sub>2</sub> | [85] [86] |
| 6 | **LRP2** | 1 |  | Renin-angiotensin homeostasis, | [87] |
| 7 | **CDK14** | 1 |  | Cell-cycle and Wnt signaling | [88] |
| 8 | **TUBB4B**<br>β-tubulin isoform | 1 |  | GJA1–PI3K/AKT/KLF4<br>pathway | [89] |
| 9 | **ACAN**<br>Aggrecan | 1 |  | Promote VSMC apoptosis<br>Plaque instability | [90] |

## Conclusion

This WGS pilot dataset provides a genomic landscape for evaluating molecular drivers of early-onset cardiovascular disease and constitutes a valuable starting point for large scale study to validate these findings. It is conceivable to point out a single disease framework for PCAD: an endothelial–immune–metabolic remodeling axis that amplifies lipid retention, vascular inflammation, smooth-muscle reprogramming, and thrombosis. Within this framework, several loci could potentially emerge as pathway anchors for premature atherosclerosis leading to MI at a young age. This data provides insights into WGS as a methodology to investigate genetic architecture atherosclerosis at young age. These genomic findings described in this paper are preliminary and hypothesis-generating, derived from a small sample size intended to demonstrate feasibility and establish analytic workflows. The candidate loci and pathways require replication in larger, adequately powered cohorts. A larger sample study is underway to provide the statistical power needed to validate these signals.

## Data Availability

All data produced in the present work are contained in the manuscript

## Modified: 11.8.2025.IAC

